# Epidemiology of *Schistosoma mansoni* infection in Ituri Province, North-eastern Democratic Republic of the Congo

**DOI:** 10.1101/2020.08.10.20171090

**Authors:** Maurice M Nigo, Peter Odermatt, Georgette B. Salieb-Beugelaar, Oleksii Morozov, Manuel Battegay, Patrick R Hunziker

## Abstract

**Background:** Schistosomiasis, caused by *Schistosoma mansoni*, is of great significance to public health in sub–Saharan Africa. In the Democratic Republic of Congo (DRC), information on the burden of *S. mansoni* infection is scarce, which hinders the implementation of adequate control measures. We assessed the geographical distribution of *S. mansoni* infection across Ituri province in north-eastern DRC and determined the prevailing risk factors.

**Methods / Principal Findings:** Two province–wide community–based studies were conducted. First, in 2016, a geographical distribution study was carried out in 46 randomly selected villages, covering 12 of the 36 health districts across Ituri. Second, in 2017, an in–depth study was conducted in 12 purposively-selected villages, across six health districts. In each study village, households were randomly selected and members, aged one year and older and present on the survey day, were enrolled. In 2016, one stool sample was collected per participant, while in 2017, several samples were collected per participant. *S. mansoni* eggs were detected using the Kato–Katz technique. The 2017 study also incorporated a point–of–care circulating cathodic *S. mansoni* antigen (POC–CCA) urine test. Household and individual questionnaires were used to collect data on demographic, socioeconomic, environmental, behavioural and knowledge risk factors.

The 2016 study included 2,131 participants, 40.0% of whom had *S. mansoni* infections. Infection prevalence in the villages ranged from 0 to 90.2%. The 2017 study included 707 participants, of whom 73.1% tested positive for *S. mansoni*. Infection prevalence ranged from 52.8 to 95.0 % across the health districts visited. In general, infection prevalence increased from north to south and from west to east. Exposure to the waters of Lake Albert and the villages’ altitude above sea level were associated with the distribution.

Both men and women had the same infection risk (odds ratio [OR] 1.2, 95% confidence interval [CI] 0.82-1.76). Infection prevalence and intensity peaked in the age groups between 10 and 29 years. Preschool children were highly infected (62.3%). Key risk factors were poor housing structure (OR 2.1, 95% CI 1.02-4.35), close proximity to water bodies (OR 1.72, 95% CI 1.1-2.49), long–term residence in a community (OR 1.41, 95% CI 1.11-1.79), lack of latrine in the household (OR 2.00, 95% CI 1.11-3.60), and swimming (OR 2.53, 95% CI 1.20-5.32) and washing (OR 1.75, 95% CI 1.10-2.78) in local water bodies. A family history of schistosomiasis (OR 0.52, 95%, 95% CI 0.29-0.94) and knowledge of praziquantel treatment (OR 0.33, 95% CI 0.16-0.69) were protective risk factors, while prevention knowledge (OR 2.35, 95% CI 1.36-4.08) was associated with increased infection risk.

**Conclusions/Significance:** Our results confirm high endemicity of *S. mansoni* in Ituri province, DRC. Both the prevalence and intensity of infection, and its relationship with the prevailing socioeconomic, environmental, and behavioural risk factors indicate intense exposure and alarming transmission levels. The study findings warrant control interventions that pay particular attention to high–risk communities and population groups, including preschool children.

**Author Summary:** Intestinal schistosomiasis threatens many people in the tropical world, particularly those in Sub–Saharan Africa. Information on schistosomiasis in the Democratic Republic of Congo (DRC) is very scarce, which is a major barrier to planning and implementing efficient control programmes.

We conducted two community–based studies with the objective of assessing the geographical distribution of *S. mansoni* infection across the Ituri province in north-eastern DRC and determining the prevailing risk factors. In 2016, a geographical distribution study was carried out in 46 randomly selected villages across the province. In 2017, an in–depth study was conducted in 12 purposively selected villages. In both studies, households were randomly selected and members, aged one year and older and present on the survey day, were enrolled. In 2016, one stool sample was examined per participant, whereas several stool samples were examined for each participant in 2017. *S. mansoni* eggs were detected using the Kato–Katz technique. The 2017 study also included a point–of–care circulating cathodic *S. mansoni* antigen (POC–CCA) urine test. Household and individual questionnaires were used to collect data on demographic, socioeconomic, environmental, behavioural and knowledge risk factors. The 2016 study included 2,131 participants, 40.0% of whom had *S. mansoni* infections. The 2017 study included 707 participants, of whom 73.1% tested positive for *S. mansoni*. In general, infection prevalence increased from north to south and from west to east. Exposure to the waters of Lake Albert and villages’ altitude above sea level were main drivers of the distribution. A risk factor analysis revealed that both men and women had the same infection risk. Infection prevalence and intensity peaked in the age groups between 10 and 29 years. Preschool children were highly infected (62.3%). We identified the main risk factors to be poor housing structure, proximity to water bodies, long–term residence in a community, lack of latrine in the household, and swimming and washing in local water bodies. A family history of schistosomiasis and knowledge of praziquantel treatment were protective risk factors, while prevention knowledge was associated with increased infection risk. Our results confirm that *S. mansoni* is highly endemic in Ituri province, DRC. Both infection prevalence and intensity, and its relationship with the prevailing socioeconomic, environmental, and behavioural risk factors indicate intense exposure and alarming transmission. Control interventions are warranted and should pay attention to high–risk communities and population groups, including preschool children.

## Introduction

Schistosomiasis is widespread in sub-Saharan Africa (SSA) where it constitutes major health problem. Human infection is caused by six species of a trematode of the genus *Schistosoma*. Schistosomiasis is transmitted by the eggs of the parasite present in stools and/or urine of infected people, which contaminate the nearby water, then freshwater snails from which infective larvae are released and penetrate the skin of people during contact with infested water. The disease is multifactorial. its transmission is very focal and lies on environmental, parasitic, vector and host factors, including socio-economic and behavioural factors amongst them the lack of proper and adequate sanitation, safe water, and hygiene (WASH). It represents a major cause of global disability, morbidity, and mortality in the affected regions [1, 2]. Recent estimates suggest that nearly 800 million people are at risk for schistosomiasis, while 240 million are infected worldwide [3]. More than 90% of all infected people live in sub–Saharan Africa [4]. The main control strategy is the population-based preventive chemotherapy (PCT) using praziquantel (PZQ) [5].

In SSA, four species of *Schistosoma* including *Schistosoma mansoni* which causes intestinal schistosomiasis, *S. haematobium*, the agent of genitourinary schistosomiasis, *S. intercalatum* and *S. guineensis* [6].

Democratic Republic of Congo (DRC) has the third highest reported number of cases in SSA (15 million), just after Nigeria (29 million) and United Republic of Tanzania (19 million) [7]. In DRC, three species of *Schistosoma* are reported to be present. *S. mansoni, S. haematobium*, and *S. intercalatum. S. mansoni* is abundant in the eastern region of the country along the shores of the great lakes, and in the western region, whereas *S. haematobium* is present in the central and south-eastern regions, and *S. intercalatum* in the central-northern regions of the country [8].

Intestinal schistosomiasis is one of the major neglected tropical and poverty–related diseases. *S. mansoni* thrives in tropical and sub–tropical regions with poor sanitation conditions [1, 4, 9]. *S. mansoni* alone threatens about 393 million people in Africa, the Middle East, Brazil, Venezuela, Suriname and the Caribbean, and about 54 million are infected [10]. Children bear the highest burden of infection because of their inadequate hygiene and frequent contact with infected waters [11].

In the DRC, the current burden of this neglected tropical disease (NTD) is unknown [12]. The DRC suffers from a recent history of war and ongoing tribal and armed conflicts, the results of which have led to economic deterioration, severe poverty, and badly functioning health services. Data from the available large surveys are more than twenty–years old [8, 13]. Existing publications reporting on schistosome infection focus on different topics [14] or mostly describe epidemiological studies carried out in a few areas of the country [8], such as Kinshasa [15-18], the capital city; and in the provinces of Kongo Central [19-21], Bandundu [22] Kwilu [23, 24], Kasaï,Central, Kasaï Oriental [13], Maniema [25-30], South–Kivu [31], Katanga [32, 33] and Haut-Uele province [34].

Publications relating to the north-eastern provinces are scarce [26, 31, 34] or date back to the colonial period [8, 30]. In 2012, the Ministry of Health (MOH) launched a national survey of NTDs and a new strategic plan was elaborated in 2016 [35, 36]. Although a national plan against NTDs, including schistosomiasis, has been implemented by the Ministry of Health, the intervention strategy is based on outdated prevalence data in many regions. Ituri province in north-eastern DRC is one such case, in which the strategy applied is for moderate–risk settings (10 – 49%), yet it appears to be a high–risk area. In this situation, the large number of infected and untreated people will contribute to perpetuating the transmission as long as treatment coverage remains inadequate [37]. Therefore, it is urgent to update our knowledge regarding the distribution of the intestinal schistosomiasis to undertake appropriate intervention and assessment.

The aim of this work was (i) to characterize the prevalence and intensity of *S. mansoni* infection and (ii) to identify key risk factors of *S. mansoni* infection in Ituri province, north-eastern DRC.

## Materials and Methods

### Ethics statement

This study was approved by the Swiss Ethical Commission (Ref. No. UBE–15/78) and by the University of Kisangani’s Research Ethical Commission, (Ref No: CER/003/GEAK/2016). Research authorization was granted by the Nyankunde Higher Institute of Medical Techniques (Ref No 70/ISTM–N/SGAC/2017), Bunia, DRC. Permission for field work was obtained from Ituri Provincial Health Division (Ref. 054/433/DPS/IT/06/2016 and Ref. 054/472/DPS/IT/06/2017) and from all relevant health districts. Prior to enrolment, the study objectives and procedures were explained to the participant in the local language. All participant questions were answered. Written informed consent was obtained from all study participants aged 15 years and above. Parents or legal guardians signed consent forms for participants aged 1-14 years. All participants diagnosed with *S. mansoni* infection were treated with praziquantel (40mg/kg) [38]. All participants received Mebendazole (500mg, single dose, Vermox®) for general deworming, in accordance with the DRC national deworming guidelines.

### Study area

Ituri province is situated in north-eastern DRC and has a surface area of 65,658 km^2^ (Figure 1). It is divided into counties – territories – (Aru, Mahagi, Djugu, Irumu, and Mambasa) and 36 health districts (See Figures S2a and S2b). Ituri province is heavily irrigated by natural water streams. It is characterized by a great geographical and demographic variability (Figure S1). Aru territory, in the north, is a plateau area of about 1,100 m above sea level and covered with a half-wooded and half-grassy savannah and forest galleries. Next is Mahagi territory, showing almost the same vegetation characteristics as that of Aru. However, the altitude peaks at around 1,800 m followed by a steep slope which descends to lake Albert to the east. Djugu is the high hill (up to 2,300 m above sea level in the Blue Mountains chain) territory of the Ituri province. The largest area is covered with grassy savannah and few forest galleries in the west. The mountains slope steeply towards Lake Albert to the east. Irumu territory shows the same characteristics than Djugu in the east whereas it is covered with dense forest in the west. Mambasa territory is a lowland region totally covered with equatorial dense forest. The health districts are distributed among territories according to the population density. Thus, different district health centres and communities are in different ecological settings. In all the health districts, curative and preventive activities such as vaccination, infants’ and prenatal clinics, deliveries, and human immunodeficiency virus (HIV) testing are currently organized. Some health districts do tuberculosis and leprosy care and control activities. Laboratory diagnostic activities for current diseases such as malaria and soil-transmitted helminths are available in all the district general hospitals. However, many health centres lack diagnostic tools and/or personnel. Praziquantel is available in all the health districts bordering the lake. Neglected tropical diseases (NTDs) drugs are sometimes provided in some health districts by the provincial branch of the national control programme. About 5.3 to 9.0 [39] million people of Sudanese, Nilotic, Bantu, Nilo-Hamite, and Pygmy ethnicities live in Ituri province (Figure S3). About 91% of the population live in 45.0% of the northern and eastern areas of the province. As unemployment is a major scourge in DRC, the majority of the population, both in rural and urban areas, is mainly engaged in subsistence farming. Some, however, raise small and large livestock. Young people are involved in the artisanal exploitation of minerals such as gold, diamonds and coltan. Cattle breeding is mainly practiced in Djugu, Irumu, Mahagi and Aru territories. Fishing is the main activity of people living by the lake and timber harvesting for commercial and domestic purposes is widely practised in the forested areas of the province, particularly in the Mambasa territory. These occupations coupled with the general lack of safe water put the population in frequent contact with contaminated waters of streams and lake which increases the exposure to water-transmitted parasites.

For more than two decades, Ituri province has been subject to war, turmoil, and social conflict. The socioeconomic situation in Ituri province is challenging, with a high degree poverty and precarity. The DRC ranked 176/188 in the Human Development Index in 2017 [40]. In 2011, a Water and Sanitation Program (WSP) strategic overview estimated that 50 million Congolese (75.0%) did not have access to safe water, while approximately 80-90% did not have access to improved sanitation [41]. Likewise, the UNICEF/WHO [42] 2017 database showed that in 2015, 84% of DRC’s rural population had no hygiene facility, 45.3% had unimproved sanitation, 10.2% resorted to open defecation, 53% used unimproved water sources, and 16.0% used surface water. According to data from the Food and Agricultural Organization (FAO), Ituri province measures 0.4 to 0.45 on the normalized difference vegetation index (NDVI) [43] (Figure S1). Since 2018, Ituri province is experiencing its first Ebola epidemic and as the COVID-19 pandemic emerges currently as a huge global problem, this will make the situation even more complicated, not to mention all the people that will get ill and die.

**Figure 1.**
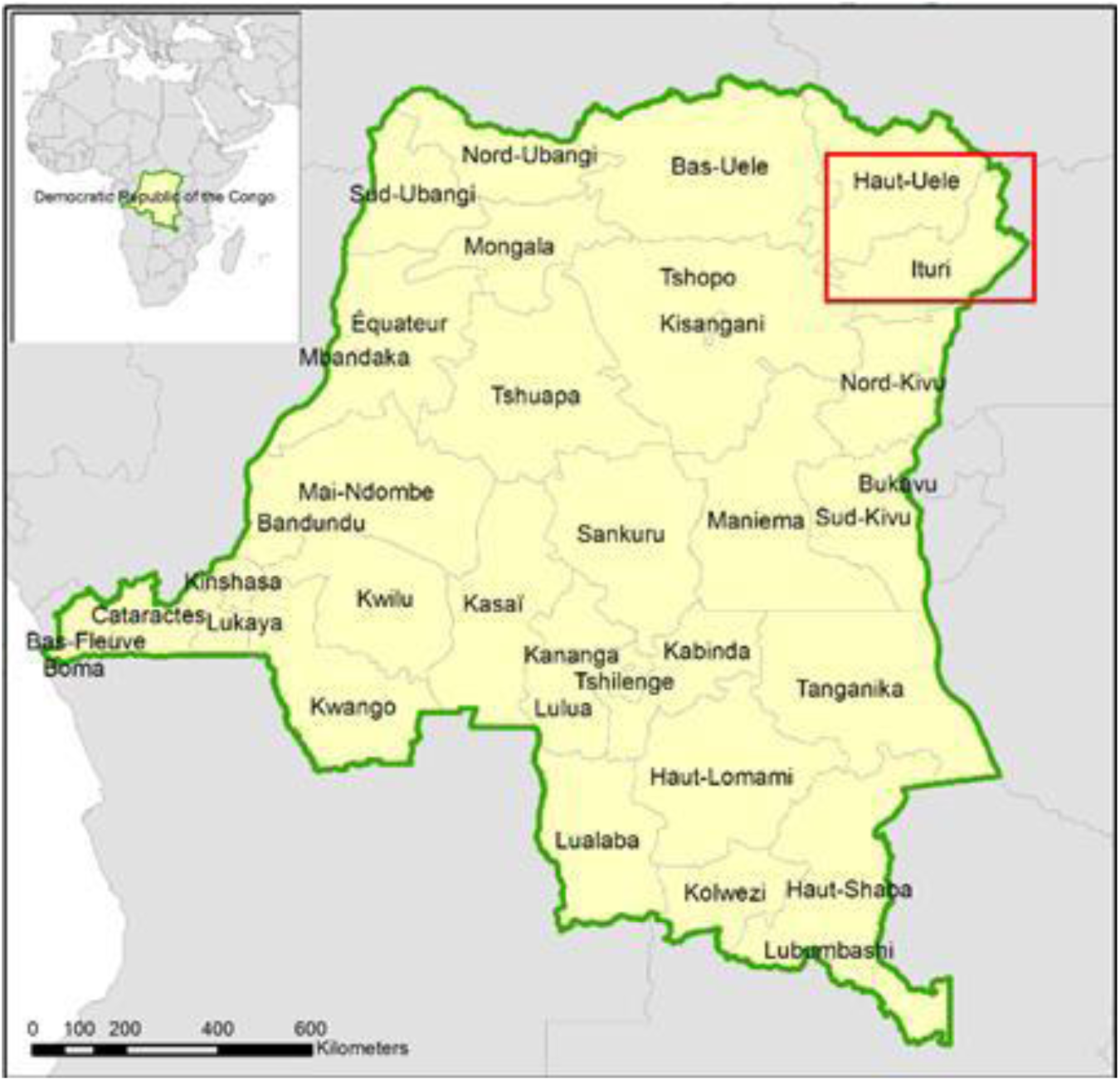
Study site: Ituri province among the 26 provinces of DRC. Map of Democratic Republic of the Congo in the centre of Africa, and Ituri province, situated among the 26 provinces, in the north-eastern part of the country.

### Study design and population

Two community–based studies were carried out across Ituri province during the long school holidays corresponding to the second dry season of the year (June to July) so as to include as much as possible children and their families in the study. First, in 2016, a cross–sectional study was carried out in 46 randomly selected villages to assess the geographical distribution of *S. mansoni* infection and infection intensity. Second, in 2017, an in–depth, cross–sectional study was conducted in 12 purposively selected villages. For both studies, people aged one year and older were eligible to participate.

The 2016 cross–sectional study used a two–stage random sampling procedure. First, 12 health districts were randomly selected across the province. Second, two to five villages were randomly selected per health district, proportional to the density of the population. Fifty to 100 participants were enrolled in each village.

For the 2017 in–depth study, twelve health districts with high *S. mansoni* prevalence were purposively selected. Three of the selected health districts could not be visited due to security concerns. In each of the remaining nine health districts, two to five villages were randomly selected. In each village, 10 to 30 households were randomly selected. In total, 144 households were visited. In each household, all members present on the survey date (aged one year and older) were invited to participate.

The chosen villages (communities) in 2016 and 2017 belong to specific health areas and in each health area there is a health centre that meets the criteria of geographical accessibility.

In both studies, unique reference codes were assigned to each study participant. Assuming a mean schistosomiasis prevalence of 50.0% in this population, we calculated that 100 to 150 individuals should be included per health district, for each study.

### Assessment of *S. mansoni* infection

In 2016, study participants provided one stool sample from which *S. mansoni* could be diagnosed using a duplicate Kato–Katz test (2 smears per stool) [44]. One hour after collection, stool specimens were transported to the local laboratory for examination. The smears were allowed to clear for half an hour before being read. The number of eggs detected on the smear was recorded for each helminth species separately.

Uurine samples were requested from 20.0% randomly selected participants of each village. The centrifugation pellets of these urine samples were microscopically examined (40X magnifier objectives). Before discarding the urine samples, strip tests were also performed (Combina™ 10 M, Human Diagnostics Uganda).

In 2017, participants provided stool samples on five consecutive survey days to assess the compliance of the participants to provide multiple samples, but also to evaluate the improvement of the sensitivity of the Kato-Katz technique according to the number of stool samples examined. On the last day, participants were asked to provide a urine sample as well. Both stool and urine samples were transported to the local laboratory for processing within an hour of collection. A duplicate Kato–Katz test [44] was performed (2 smears per stool) on each stool sample. Again, the number of eggs detected on the smear was recorded for each helminth species separately. In addition, an *S. mansoni* –based point–of–care cathodic circulating antigen (POC–CCA) test [45] was carried out on the urine samples. We combined the Kato–Katz and POC–CCA technique to overcome the weakness and shortcomings of the former technique, through the strength and quality of the latter, and *vice versa*, and to compare the testing performance of the two approaches in our population.

For quality control, about 10-30% randomly selected Kato–Katz slides were re-read by the principal investigator. Urine POC–CCA test trace and weakly positive results were discussed by the laboratory team and the principal investigator.

### Questionnaire data

In 2016, demographic information was collected with a short questionnaire addressed to each study participant, while both a household and an individual questionnaire was used in 2017. The head of household answered the household questionnaire. It included questions regarding the number of household members, the availability and quality of sanitation (latrine), the source and type of water used, the building material of the house, household goods, estimated monthly income and the existence of mass drug administration (MDA) for the benefit of the villagers. The individual questionnaire addressed demographic details (age, sex, tribal group, religion, main occupation, education, and usual defecation place), knowledge of schistosomiasis (disease, transmission, prevention, and treatment) and potential risk factors (exposure to water, socioeconomic status, time of residence, lack of sanitation and safe drinking water, etc.).

### Data management

In both studies, data were entered in Excel and cross checked with the source data. Data management and data analysis were performed with Stata, version 14.2 (Stata Corp LP; College Station, USA). Age groups were defined as follows: (i) 1-4 years, (ii) 5-9 years, (iii) 10-14 years, (iv) 15-19 years, (v) 20-29 years, (vi) 30-39 years, (vii) 40-49 years, (viii) ≥50 years to highlight as much as possible the level of infection in infants, children, younger and older adolescents, as well as young adults, adults, and older people. Body mass index (BMI) was calculated and four BMI categories were established: underweight (<18.5 kg/m^2^), normal weight (18.5-24.9 kg/m^2^), overweight (25.0-29.9 kg/m^2^), and obese ≥30 kg/m^2^). Using the arithmetic mean of the egg positive stool samples, *S. mansoni* infection intensity (eggs per gram [EPG]) was categorized as light (1-99 EPG), moderate (100-399 EPG), and heavy (≥400 EPG) [44, 46].

**Figure 2: Study field procedures**

Enrolment of study participants, administration of household and individual questionnaires, anthropometric measurements, and collection of stool samples.

### Statistical analysis

Descriptive statistics (means and frequencies) were used to summarize continuous and categorical variables, respectively. The chi–square test (χ^2^) and Fisher exact test were used to compare proportions. An univariable logistic regression analysis was performed to associate *S. mansoni* infection (outcome) with potential risk factors (predictors), such as demographic, geographical, behavioural, and socioeconomic variables. Co–variables exhibiting an association at a significance level of at least 20%, as determined by the likelihood ratio test (LRT), were included in the multivariable logistic regression models. Odds ratios (OR) and 95% confidence intervals (95% CI) were calculated. To visualize prevalence rates at health district and village levels, a twoway scatter bar command, which displays numeric (y, x) data as histogram-like bars, was performed using the vertical bar plot option in STATA. Bars were drawn at the specified xvar values (health districts and villages) and extended up from 0 according to the corresponding yvar values (prevalence). To explore the relationship between *S. mansoni* infection risk and age, age– and sex–prevalence curves were produced using the twoway quadratic fitted values. This command calculates the prediction for yvar (prevalence or intensity) from a linear regression of yvar on xvar and xvar^^^2 and plots the 15 resulting curve. An *S. mansoni* distribution map was made as follows: village prevalence (resulting from the Kato–Katz tests in 2016 and 2017) and geographic decimal coordinates (latitude and longitude) were entered in two adjoining columns in Excel and then plotted. A non-uniform spline interpolation was performed with MATLAB®, revealing shadows, the intensity of which were proportional to prevalence levels. Dark coloured shadows indicate high prevalence and lighter shadows (high transparency) indicate low prevalence. *P*–values below 5% were considered statistically significant.

## RESULTS

### Study population

Of the 2,322 individuals enrolled in 2016, 2,131 participants completed all examinations and were included in the final analysis (Figure 3). Participants represented 46 villages, with at least 10 participants from each village except for Ramogi village (Angumu health district), where only 6 participants were included (Table S1d). Table 1 shows the demographic characteristics of participants in the *S. mansoni* geographical distribution study. There were slightly more female (51.0%) than male participants. The mean age of participants was 22.2 years, with little difference between men and women. Children under 15 years made up almost half of all study participants. Nearly two thirds of participants (58.0%) had a normal BMI, one third (33.0%) were underweight and nine percent were overweight or obese. More than three quarters of the overweight/obese group were female.

**Figure 3.**
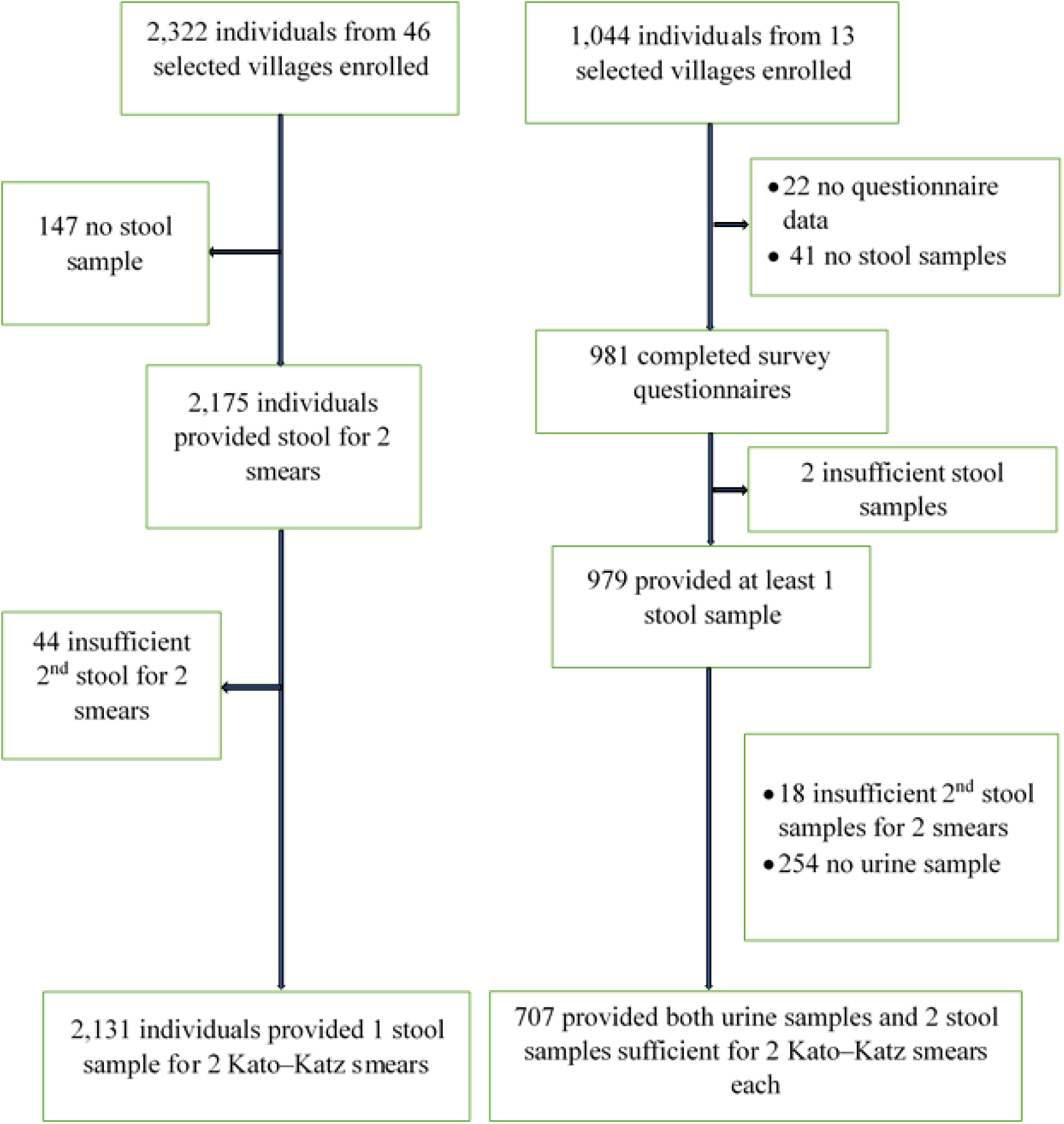
Study participants’ inclusion flowchart for the Ituri schistosomiasis surveys. Left: In 2016, geographical distribution study in 46 villages; Right: In 2017, in–depth study in 12 villages.

**Table 1.** Geographical distribution study 2016: Demographic characteristics of the participants. Cross–sectional study of geographical distribution of *S. mansoni* infections, conducted in 46 randomly-selected villages in 12 of the 36 health districts in Ituri province (n=2,131).

| Characteristics | Subgroups | Total n (%) | Female n (%) | Male n (%) |
| --- | --- | --- | --- | --- |
| Overall n (%) |  | 2,131 (100) | 1,087 (51.0) | 1,044 (49.0) |
| Mean |  |  |  |  |
| Age (years) |  | 22.2 | 22.8 | 21.6 |
| Body Mass Index (BMI) |  |  |  |  |
| Obese | (BMI≥30.0) | 35 (1.6) | 30 (85.7) | 5 (14.3) |
| Overweight | (BMI=25.0–29.9) | 157 (7.4) | 121 (77.1) | 36 (22.9) |
| Normal weight | (BMI=18.5–24.9) | 1,235 (58.0) | 634 (51.3) | 601 (48.7) |
| Underweight | (BMI<18.5) | 704 (33.0) | 302 (42.9) | 402 (57.1) |
| Children | (<18 years) | 1,107 (52.0) | 507 (45.8) | 600 (54.2) |
| Adults | (≥18 years) | 1,024 (48.0) | 580 (56.6) | 444 (43.4) |
| Age categories (years) |  |  |  |  |
| 1 – 4 |  | 26 (1.2) | 12 (46.2) | 14 (53.8) |
| 5 – 9 |  | 436 (20.5) | 202 (46.3) | 234 (53.7) |
| 10 – 14 |  | 496 (23.3) | 228 (46.0) | 268 (54.0) |
| 15 – 19 |  | 241 (11.3) | 118 (49.0) | 123 (51.0) |
| 20 – 29 |  | 337 (15.8) | 208 (61.7) | 129 (38.3) |
| 30 – 39 |  | 236 (11.7) | 132 (55.9) | 104 (44.1) |
| 40 – 49 |  | 199 (9.3) | 115 (57.8) | 84 (42.2) |
| ≥50 | | 160 (7.5) | 72 (45.0) | 88 (55.0) |
| Residence |  |  |  |  |
| Rural |  | 1 951 (91.6) | 992 (50.8) | 959 (49.2) |
| Urban |  | 180 (8.4) | 95 (52.8) | 85 (47.2) |
| Ethnic groups |  |  |  |  |
| Nilo–Hamites |  | 106 (5.0) | 48 (45.3) | 58 (54.7) |
| Bantu |  | 398 (18.7) | 222 (55.8) | 176 (44.2) |
| Nilotic |  | 753 (35.3) | 368 (48.9) | 385 (51.1) |
| Sudanese |  | 582 (27.3) | 293 (50.3) | 289 (49.7) |
| Pygmies |  | 292 (13.7) | 156 (53.4) | 136 (46.6) |

Of the 1,044 individuals enrolled in the 2017 study, 707 completed all procedures (Figure 3). Participants lived in one of 12 villages (144 households), with at least 23 participants present in each village (Table S2a). Table 2 shows the demographic characteristics of participants in the in–depth study carried out in 2017.

**Table 2.** In–depth study 2017: Demographic characteristics of the participants. Study conducted in 12 purposively selected villages in 6 of the 36 health districts in Ituri province (n=707).

| Characteristics | Subgroups | Total n (%) | Female n (%) | Male n (%) |
| --- | --- | --- | --- | --- |
| Overall n (%) |  | 707 (100) | 398 (56.3) | 309 (43.7) |
| Mean |  |  |  |  |
| Age (years) |  | 21.0 | 21.6 | 20.2 |
| Family size (no) |  | 8.6 | 8.5 | 8.6 |
| Body Mass Index (BMI) |  |  |  |  |
| Obese (BMI≥30.0) |  | 63 (8.9) | 51 (81.0) | 12 (19.0) |
| Overweight (BMI=25.0–29.9) |  | 24 (3.4) | 23 (95.8) | 1 (4.2) |
| Normal weight (BMI=18.5–24.9) |  | 271 (38.3) | 158 (58.3) | 113 (41.7) |
| Underweight (BMI<18.5) |  | 349 (49.4) | 166 (47.6) | 183 (52.4) |
| Children (<18 years) |  | 406 (57.4) | 205 (50.5) | 201 (49.5) |
| Adults (≥18 years) |  | 301 (42.6) | 193 (64.1) | 108 (35.9) |
| Age categories (years) |  |  |  |  |
| 1 – 4 |  | 61 (8.6) | 29 (47.5) | 32 (52.5) |
| 5 – 9 |  | 158 (22.4) | 77 (48.7) | 81 (51.3) |
| 10 – 14 |  | 144 (20.4) | 73 (50.7) | 71 (49.3) |
| 15 – 19 |  | 68 (9.6) | 40 (58.8) | 28 (41.2) |
| 20 – 29 |  | 83 (11.7) | 67 (80.7) | 16 (19.3) |
| 30 – 39 |  | 75 (10.6) | 49 (65.3) | 26 (34.7) |
| 40 – 49 |  | 56 (7.9) | 32 (57.1) | 24 (42.9) |
| ≥50 |  | 62 (8.8) | 31 (50.0) | 31 (50.0) |
| Residence |  |  |  |  |
| Rural |  | 377 (53.3) | 216 (57.3) | 161 (42.7) |
| Urban |  | 330 (46.7) | 182 (55.2) | 148 (44.8) |
| Ethnic groups |  |  |  |  |
| Nilo–Hamites |  | 160 (22.6) | 98 (61.3) | 62 (38.7) |
| Bantu |  | 312 (44.1) | 179 (57.4) | 133 (42.6) |
| Nilotic |  | 166 (23.5) | 88 (53.0) | 78 (47.0) |
| Sudanese |  | 69 (9.8) | 33 (47.8) | 36 (52.2) |

Female participants (56.3%) outnumbered male participants. More than half of the participants (51.4%) were younger than 15 years. Almost half of the participants (49.4%) were underweight, 38.3% had a normal weight, and 12.3% were overweight or obese. More than 80% of the overweight/obese group were female.

### *Schistosoma mansoni* infection prevalence and intensity

Table S1a summarizes results of the two surveys for the different diagnostic approaches. The global results are shown as prevalence and intensity of infection with the 95% confidence intervals (95% CI). Detailed analysis of the results by health districts, villages, sex, age groups, residence, ethnic groups, and altitude are then shown through Tables S1b-S1d and S2a-S2i.

## Prevalence

In 2016, the overall *S. mansoni* average prevalence measured by Kato–Katz was 40.0% (37.9-42.1) (Tables S1a) ranging from 3.9% to 80.2% across the 12 health districts. Detailed results showed significant variation of infection prevalence between sex, age categories, ethnic groups, altitude levels, villages, and health districts. Infected individuals were found in 43 of the 46 (93.5%) of the investigated villages. Male study participants (42.6%) were more frequently infected than females (37.4%) (p=0.015). The difference in the prevalence of infection across the eight age groups (Table S1b) and in both sexes (Table S1c) was highly significant (p<0.001). The prevalence in the youngest age group (1-4 years) was a quite low (7.7%), being 8.3% and 7.1% in girls and boys, respectively. This figure rose clearly in subsequent age groups to reach a maximum of 50.2% in the 15-19 years, being 45.8% in females and 54.5% in males. Then, the prevalence declined slightly after this peak but remained generally high throughout.

Among the twelve health districts surveyed, *S. mansoni* infection was highest in the health districts of Nyarambe (80.2%), Tchomia (73.7%), and Angumu (70.2%) all situated at the shore of Lake Albert. They are followed by Lolwa (52.5%), Komanda (50.0%), Nyankunde (49.2%), and Bunia (33.9%) situated in the central and south-western regions of the province. For the rest of the health districts situated in the high-hill region (Bambu, Rethy, Logo) or in the northern region (Adi and Laybo), the overall prevalence was less than 10.0%. The difference in the prevalence of infection across the twelve health districts was highly significant (p<0.001) (Table S1d and Figure 4). The difference in the prevalence of infection according to the altitude above the sea level – high (>1,800 m), middle (1,000-1,800 m), and low (<1,000 m) – was highly significant in both sexes (p<0.001) (Table S1b and Figure 8). However, the difference between rural areas (40.5%) and urban area (33.9%) (p=0.081), and both among females (p=0.568) and males (p=0.060), was not significant. At the village level, infection prevalence varied widely but was mostly consistent with the health district mean value. In three of the 46 villages, no *S. mansoni* infection was detected. These villages were Upepeni and Ombanya, in the Adi and Laybo health districts, respectively (northern Ituri province), and Fundi, in the Rethy health district (located in the hills at about 1,920 m above sea level). Villages along the shore of Lake Albert had prevalence rates over 70%, with the highest prevalence rate reaching 90.2% in Kolokoto (at the Ugandan border), in the Nyarambe health district (Table S1d and Figure 5).

**Figure 4.**
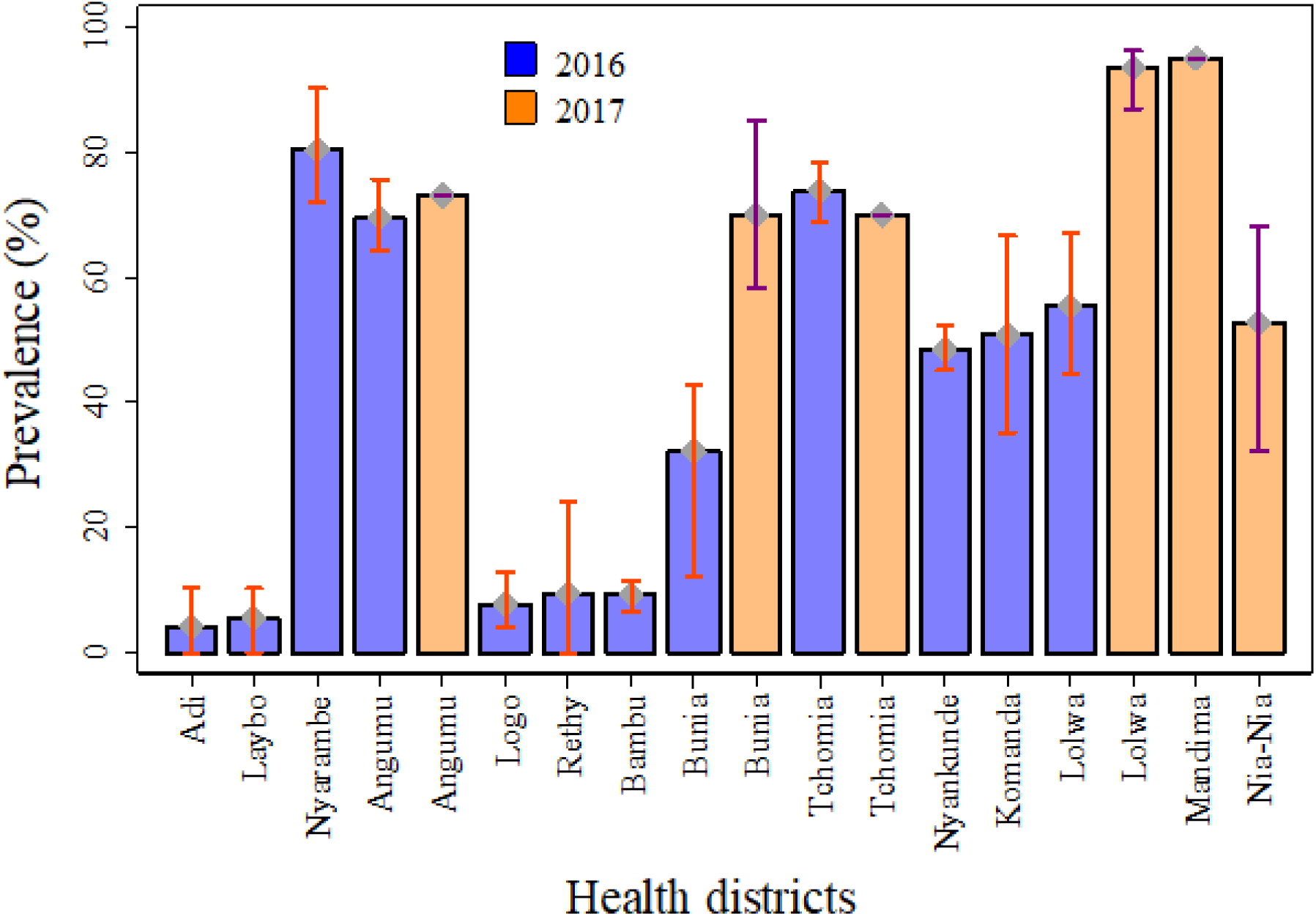
*S. mansoni* prevalence by health district in 2016 and 2017. Blue bars: 2016 study (46 study villages across 12 health districts), Kato–Katz test (two smears) of one stool sample. Orange bars: 2017 study (12 study villages across 6 health districts), Kato–Katz test of two stool samples (four smears) and one POC–CCA urine test.

**Figure 5.**
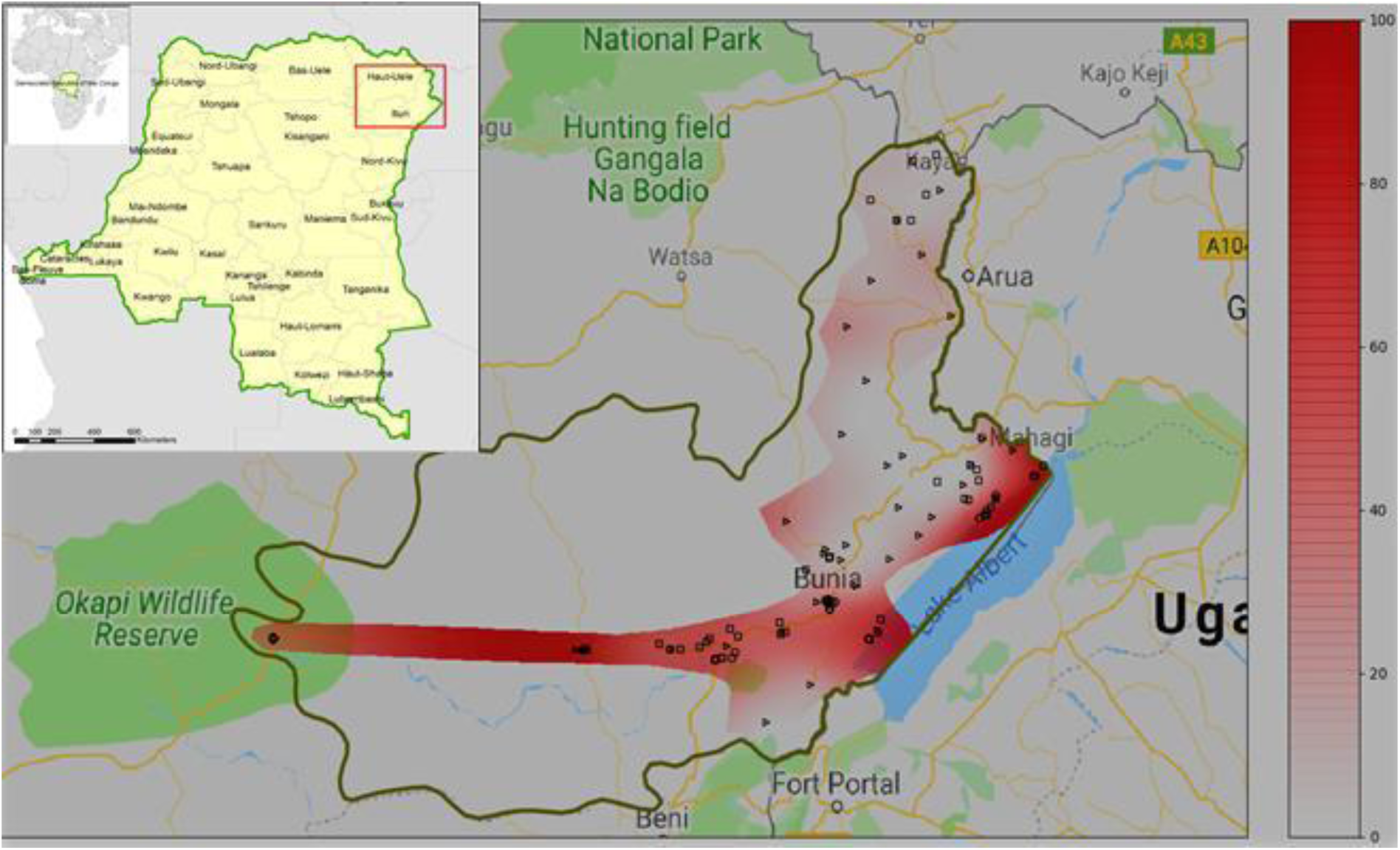
Map of Ituri province with estimated *S. mansoni* prevalence. Estimated *S. mansoni* prevalence using non-uniform spline interpolation. Intensity of red shadows is proportional to prevalence levels. Dots indicate studied villages. Areas outside of the red shadow were inaccessible, dense tropical forest and sparsely populated.

As for the ethnic groups, the prevalence of S, mansoni infection was highest in Pygmies (50.3%), followed by Bantu (48.0%), Nilotic (47.4%), and Nilo-Hamite (47.2%). The Sudanese ethnic group had the lowest (18.4%) (Table S1b), and all these differences were consistent with sex (Table S1c) and were statistically significant (*p*<0.001).

In 2017, the overall *S. mansoni* prevalence was 38.5% with one stool sample 55.0% with two stool samples examined by Kato–Katz test alone. The prevalence rose to 62.5% with one POC-CCA test alone and to 73.1% after combining the Kato–Katz results of two stool samples with one POC–CCA test findings. All the results were consistent in both sexes (Tables S2b-S2e, and S2g-S2i). In this section, we only took in account results from the combining diagnostic approach (2KK+POC–CCA) which appeared more relevant in our viewpoint (Table S2g). Herein also, detailed results showed significant variation of infection prevalence between health districts, villages (Table S2a), and age categories (Table S2g) (*p*<0.001). Infected individuals were found in all the 12 (100%) investigated villages. Males (74.1%) were slightly more infected than females (72.4%) (*p*=0.603). The difference in the prevalence of infection across the eight age groups was highly significant (p<0.001). This difference was observed both among female (*p*=0.030) and male (*p*=0.001) participants (Table S2i). The prevalence in the youngest age group (1-4 years) was high (62.3%), being 62.1% and 62.5% in girls and boys, respectively. This figure rose sharply in subsequent age groups to reach a maximum of 86.1% in the 10-14 years, being 84.9% in girls and 87.3% in boys. In males however, the peak reached 93.8% in 20-29 years age category. Then, the prevalence declined after these peaks but remained generally high throughout (Figure 4.

*S. mansoni* infection was high in all the six health districts surveyed, varying between 70.0-95% among the health districts of Tchomia, Bunia, Angumu, Lolwa, and Mandima. The health district of Nia-Nia, located in the extreme southwest of the Ituri province, had the lowest prevalence (52.8%). The difference in the prevalence of infection across the six health districts was highly significant (*p*<0.001) (Table S2a). The differences in the prevalence of infection according to the altitude above the sea level and rural/urban areas were not significant (*p*=0.214) both among females (*p*=0.544) and males (*p*=0.419). At village level, prevalence varied around the health district mean value (Figure 4). The lowest prevalence rate (32.3%) was found in Bankoko, Nia-Nia health district, while the highest rates (over 90%) were all found in the forest region in the southern part of the province (i.e., Mandima, 95.0% and Pekele, 96.2%, in Mandima and Lolwa health districts, respectively).

As regards ethnic groups, Pygmies could not be included in the analysis. The prevalence of *S, mansoni* infection was highest in Sudanese (85.5%), followed by Bantu (73.4%), Nilotic (71.7%), and Nilo-Hamite (68.8%). The differences were not statistically significant (*p*=0.068) and were consistent with sex (*p*=0.208) and (*p*<0.350) among females and males, respectively (Table S2g).

Comparing the prevalence of *S. mansoni* infection at the health district level (Figure 4), there does not appear to be a great difference between 2016 and 2017 rates for Angumu (70.2% and 73.1%) or for Tchomia (73.7% and 70.0%), both of which are situated on the shore of Lake Albert. However, the difference between 2016 and 2017 rates is discernible for Bunia (33.9% and 70.9%), in the central Plateau region and for Lolwa (52.5% and 93.4%), in the southern forest–covered region of Ituri province.

### Intensity of infection

The arithmetic mean intensity of *S. mansoni* infection in the twelve health districts investigated in 2016 and in the six in 2017 are shown in Tables S1d and S2a, respectively. In 2016, the egg count for S. mansoni ranged from 0 to 14,424 epg, and from 0 to 5,472 epg in 2017. The overall arithmetic mean infection intensities were 207.4 epg in 2016, and in 2017, 100.9 epg using 1KK (Table S2b) and 104.1 epg using 2KK (Table S2d), respectively.

In 2016, six health districts including Nyarambe (928.6 epg), Tchomia (640.1 epg), Angumu (392.3 epg), both located along the shore of Lake Albert, and those of Lolwa (199.3 epg), Komanda (188.9 epg), and Nyankunde (133.4 epg), located in the central and south-western areas, were most heavily infected with *S. mansoni*. The six other health districts had light infections (<100 epg), with 0.7-10.0% of moderate and 0.0-2.1% of heavy *S. mansoni* infections (Table S1d). The proportion of males with heavy infection intensity (13.8%) was higher compared to that of females (9.7%) (*p*=0.013) (Table S1b). The age distribution of *S. mansoni* infection prevalence and intensity is shown in Figure 6. In fact, the highest egg counts were found among boys of the age groups of 10-14 years (14,424 epg), 15-19 years (13,248 epg), whereas girls of the age groups of 5-9 years (10,680 epg) and 10-14 years (8,376 epg) bore the heaviest *S. mansoni* infection intensities. The difference in the intensity of infection across the eight age groups was highly significant in both sexes (*p*<0.001). The figure of infection intensity follows that of the prevalence, rising sharply to peak in 10-19 years age group and declining with the increasing age, but remaining high into the older age. As for the ethnic groups, the heaviest mean intensities in males and females were as follows: among Nilotic: 430.6 and 318.8 epg, Pygmies: 232.3 and 192.6 epg, Bantu: 185.7 and 109.9 epg, and among Nilo-Hamite: 125.0 and 162.0 epg, respectively. The lowest infection intensity was found among Sudanese: 62.6 and 23.7 epg in males and females, respectively (Tables S1b and S1c).

**Figure 6.**
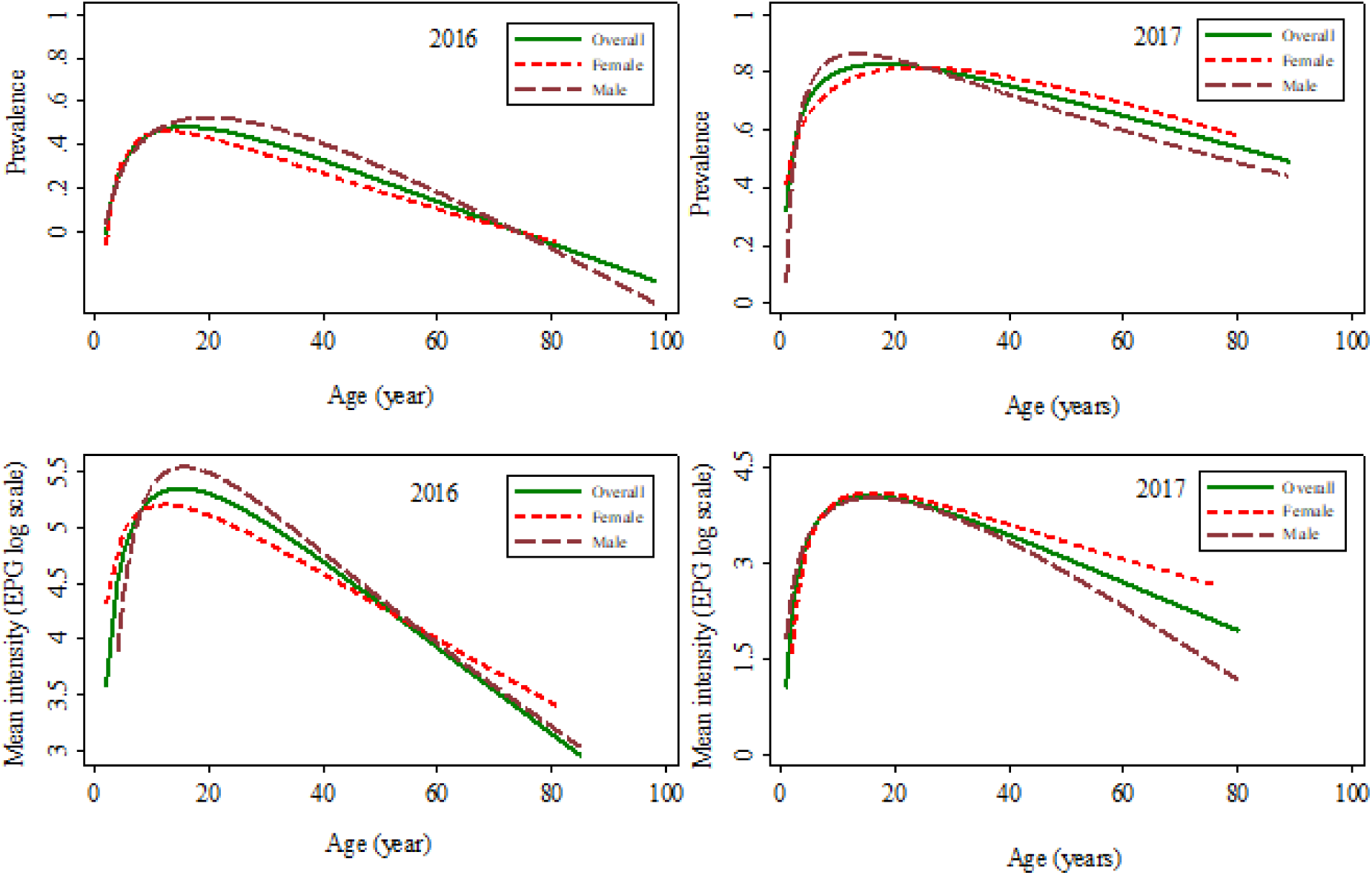
Correlation of *S. mansoni* infection prevalence and intensity with age and sex in the two studies. Top: *S. mansoni* infection prevalence by age in 2016 and 2017. Bottom: *S. mansoni* infection intensity (log scale) by age in 2016 and 2017. Lines: Green (solid): all participants; Red (dashed): female; Maroon (long dashed): male.

In 2017, the health districts of Lolwa (420.5 epg), of Mandima (131.4 epg) in the south-western forest-covered areas, and that of Tchomia (109.2 epg) in the shore of Lake Albert, were leading with heaviest infection intensities. They are followed by health districts of Angumu (83.9 epg) and Bunia (48.6 epg). The lowest infection intensity was found in Nia-Nia health district (5.8 epg). Overall, the proportion of males with heavy infection intensity (5.8%) was insignificantly lower compared to that of females (6.5%) (*p*=0.564) (Table S2b). The age distribution of *S. mansoni* infection prevalence and intensity is shown in Figure 6. In fact, the highest egg counts were found among boys of the age groups of 10-14 years (14,424 epg), 15-19 years (13,248 epg), whereas girls of the age groups of 5-9 years (10,680 epg) and 10-14 years (8,376 epg) bore the highest *S. mansoni* egg counts. The difference in the intensity of infection across the eight age groups was a quite the same as in 2016, being highly significant in both sexes (*p*<0.001) and following the figure of the infection prevalence, as described above. However, for the four remaining ethnic groups, roles have been reversed. Now, Sudanese males (281.0 epg) and females (265.4 epg) had heaviest infection intensity, followed by Bantu: 103.7 epg in males and 98.3 epg in females, respectively. Among Nilotic and Nilo-Hamite ethnic groups, males had 63.7 and 26.1 epg, and females 84.1 and 72.0 epg, respectively (Table S2d).

Figure 5 shows a map of Ituri province with the *S. mansoni* village prevalence rates observed in the 2016 and 2017 surveys. The rates were generally higher in the South than in the North and were highest in the lowlands and along the shores of Lake Albert.

The relationship between the risk of *S. mansoni* infection and age is displayed in Figure 6. In both the 2016 and 2017 studies, prevalence and intensity increased proportionally with age among both female and male participants until max. 29 years. *S. mansoni* infection prevalence peaked at around 50.0% in 2016, and at around 80.0% in 2017. While prevalence was lower in 2016, peak intensity was higher (207.3 EPG) in 2016, and lower (104.1 EPG) in 2017. Prevalence was highest among children aged 15-19 years in 2016 and decreased among older participants. Prevalence was highest among participants aged 10-14 years in 2017. Overall, the male curves peaked higher in prevalence rates in males than in females in both 2016 and in 2017. Whereas infection intensity was higher among males than among females in 2016, it was similar among males and females in 2017. The trends for different age groups, in terms of prevalence and intensity, are almost the same. However, a slight difference is observed in 2017: while infection intensity decreased abruptly for males, it remained a plateau for females.

### Correlation between *Schistosoma mansoni* infection prevalence and intensity

The relationship between *S. mansoni* infection prevalence and intensity is displayed in Figure 7. *S. mansoni* village–level infection prevalence was positively correlated with infection intensity in the 2016 and 2017 studies.

**Figure 7.**
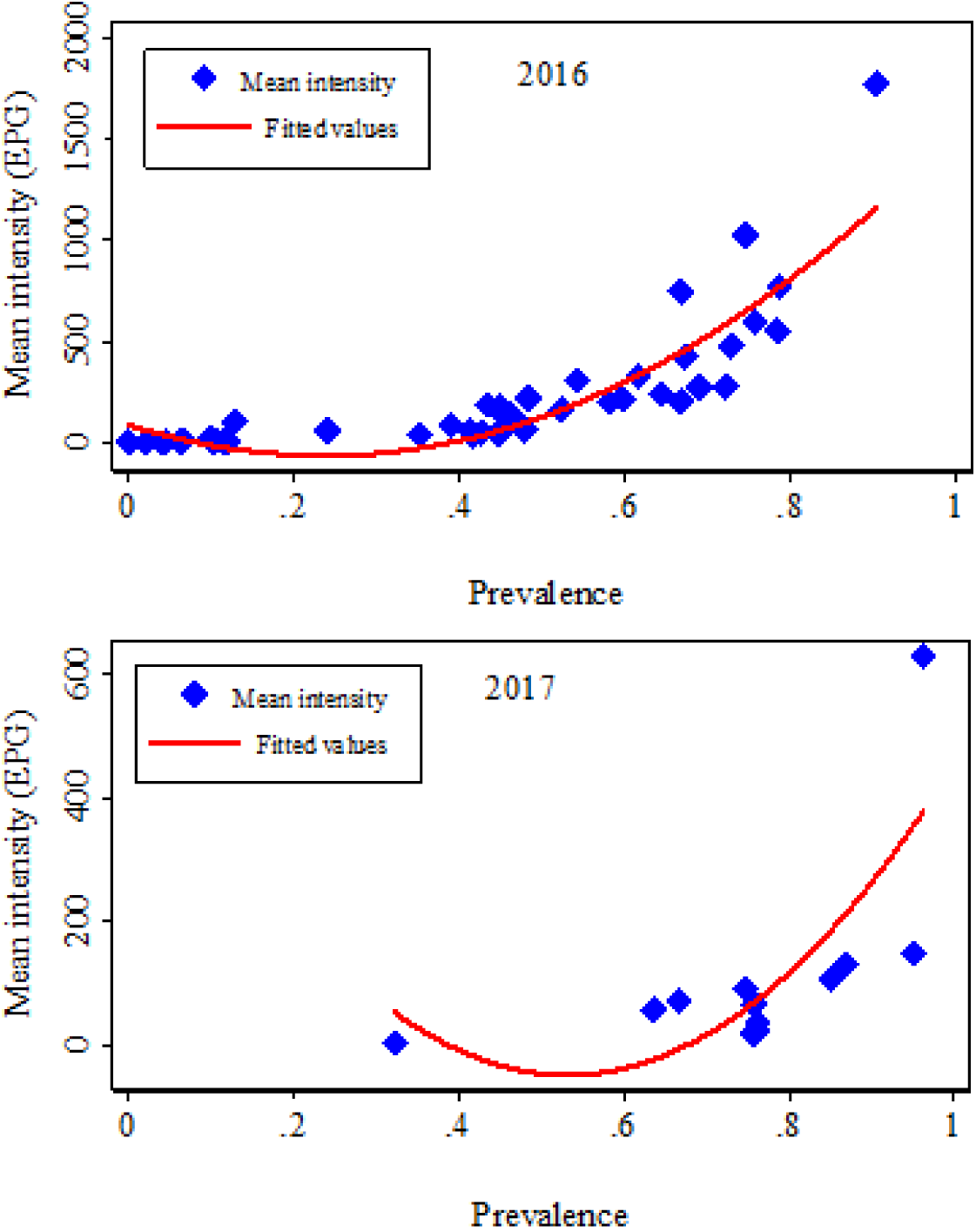
Correlation between *Schistosoma mansoni* infection prevalence and intensity at village level in the 2016 geographical study (top) and the 2017 in–depth study (bottom) All quadratic line fitted values.

A relationship was also observed between *S. mansoni* village-level infection prevalence and intensity and altitude in the 2016 study (Figure 8). The *S. mansoni* infection prevalence and intensity were negatively correlated with altitude.

**Figure 8.**
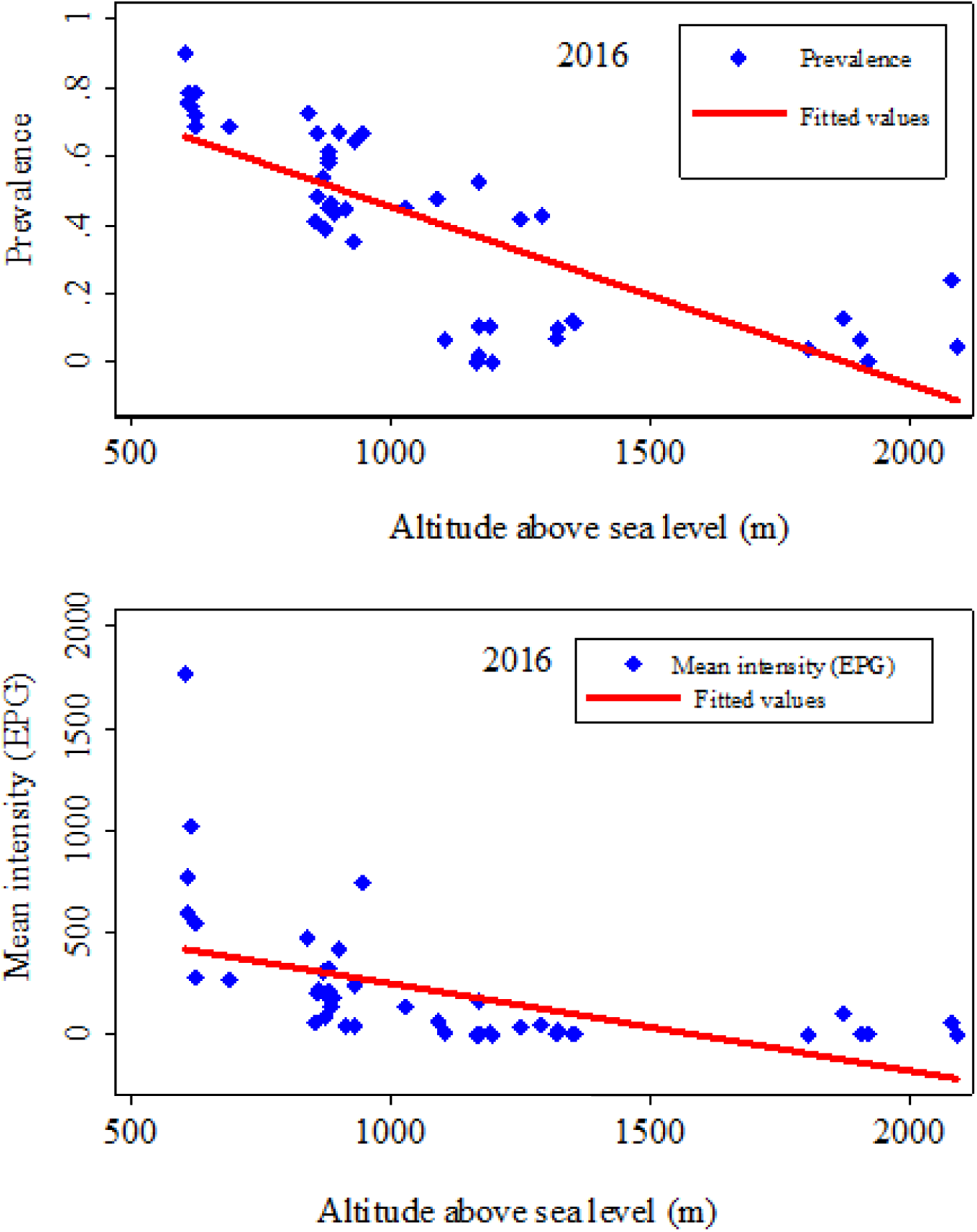
Correlation of *S. mansoni* infection prevalence and intensity with altitude (meters above sea level) in the 2016 distribution study. The figure above shows the influence of altitude on *S. mansoni* infection prevalence (top) and intensity expressed as an arithmetic mean (bottom) as observed in 2016. The higher the altitude, the lower the infection prevalence and intensity.

### Risk factors for *Schistosoma mansoni* infection

Tables S3a–S3d show the results of the univariable risk analysis for an *S. mansoni* infection with demographic, socioeconomic, environmental, behavioural, family, and individual variables. A significant increase in risk is associated with age (10-29 years), ethnicity (Sudanese), duration of residence (≥10 years), bad housing, and not owning shoes. *S. mansoni* infection was also associated with living in the health districts of Bunia, Tchomia, Angumu, Lolwa and Mandima, and with proximity of the household to a nearby body of water. The absence of a latrine in the household, washing clothing in streams, and farming were among the most high–risk behaviours. Knowledge about the use of praziquantel was found to be a protective factor against *S. mansoni* infection.

Predictive variables with a significance level of less than 20% in the univariate model were retained in the multivariate risk factor model. Gender was also included in the multivariable logistic regression model (see Table S4).

Figure 9 presents the results of the multivariable risk factor analysis. Ten of 17 variables were significantly associated with *S. mansoni* infection. Participants living in certain health districts, such as Bunia, the unique urban area; Tchomia and Angumu, at the shore of Lake Albert; and Lolwa and Mandima, at the southern and forest-covered region of the province (p<0.005) had a significantly increased infection risk. Those with poorly built households (p<0.044), or without a latrine (p=0.022) had higher odds of being infected. Water contact activities such as swimming (p=0.014) and washing clothes in streams (p=0.018) were also associated with a significant risk increase. Furthermore, infection risk increased significantly with a longer residence period (≥10 years *vs* shorter period) and closer proximity to water bodies (p=0.005). Finally, participants who had a family history of schistosomiasis (p=0.030) and those with knowledge of praziquantel treatment had a significantly lower *S. mansoni* infection risk (p=0.003).

**Figure 9. Results of multivariable logistic regression analysis for *S. mansoni* infection in 2017**

Risk analysis performed for participants from 12 villages in Ituri province (n=707). Odds ratios (OR), large red diamond and 95% confidence interval (CI), range indicated by horizontal blue lines.

## Discussion

To the best of our knowledge, this study is the first comprehensive, province-wide assessment of *S. mansoni* infection prevalence, intensity, and risk factors in Ituri since colonial times. In two studies, encompassing 14 (38.9%) of the 36 health districts in Ituri province (Figure S2b), including 56 villages and more than 2,800 participants, we firstly used Kato–Katz test alone in 2016 and, alarmed by the findings, applied a more sensitive diagnostic approach which combined both Kato–Katz and POC–CCA tests results and found a very high *S. mansoni* infection burden, revealing a major public health problem in the province.

In the 2016 study, conducted in 46 villages across 12 health districts, 40.0% of the study participants tested positive for *S. mansoni*. First, Men showed a higher infection prevalence and displayed heavier infection intensity than women and *S. mansoni* infection was evidently acquired early in the life as many children below 5 years were found infected. Both prevalence and intensity peaked in the 10 – 14 and 15 – 19 years age groups. These results may reflect, on one hand, the more frequent exposure of men to contact with water than women through fishing and farming activities, because culturally in Ituri, men are responsible for meeting the food, clothing and financial needs of their families, while women, on the other hand, are more concerned with domestic needs such as fetching water, washing clothes and dishes, caring for children and preparing food. Thus, they have less contact with water than men. In endemic areas, individuals, mainly children, spend long hours swimming, playing, bathing, or fetching water from water bodies that may contain cercariae. They also defaecate indiscriminately in the environment, causing most of the contamination and increasing their own risk to infection [47].

Second, considerable variability in infection prevalence was observed at the health district level, ranging from 3.9% in Adi to 80.2% in Nyarambe. Three villages were found free of S. mansoni infection, two in the northern side and one in the high hill region. For the two first villages, we did not find an evident reason since these villages are all built near streams that are currently used for washing and bathing. However, our results are consistent with the low *S. mansoni* infection rate in colonial times, when Aru territory prevalence was reportedly below 3.0% [8, 48]. Concerning the village of Fundi, the high altitude with generally cold temperature and water velocity would not be suitable for snails reproduction. Due to the same reason, people may also have fewer contacts with water, thereby reducing the probability to become infected [49, 50]. In general, we observed higher infection rates in the south and east of the province compared to the north and west. Data from colonial times [51, 52] and from the mid–1980s [53] showed a similar geographical pattern. Indeed, exposure to the waters of Lake Albert explain this distribution pattern to a large extent. *Biomphalaria* molluscs are abundant in the lake’s shallow waters and inhabitants of the lake–side villages intensively use the water for various activities. We also found a negative association between *S. mansoni* infection and altitude. In the colder hillside areas, *Biomphalaria* mollusc development decelerates and human water contact is less frequent, leading to reduced transmission of *S. mansoni* infection [54]. Hence, the Blue Mountains chain of Ituri contributes to a lower infection prevalence pattern in the east of the province.

Surprisingly, *S. mansoni* infection prevalence was very high in the villages situated in the southwestern forest–covered part of the province, i.e. the prevalence in Lolwa and Mandima health districts (Figure S2b) exceeded the rates recorded at the lakeshore. However, this region is situated in the lowlands and the population depends on the existing water bodies in the area.

We observed that prevalence was slightly higher in rural areas (75.1%) compared to urban areas (70.9%). Schistosomiasis particularly affects poor communities [1, 55-57] without the means to protect themselves from risky water contacts. Hence, rural communities are especially affected [58-62], but suburban and urban poor communities should also be considered [63-65]. Thus, it is not surprising that the participants living in Ngezi village in Bunia city had a very high *S. mansoni* infection burden, comparable to villages on the shore of Lake Albert. In fact, Ngezi village is situated along the Nyamukau and Ngezi rivers, where adequate water supply and sanitation is lacking.

Infection prevalence was lowest in the health district of Nia–Nia, in the southwestern corner of the province. The remoteness of the villages and the scarcity of water explain the low infection rate. Furthermore, the intermediate host snail *Biomphalaria alexandrina stanleyi* described in this region [66] is thought to be less effective in transmitting *S. mansoni*.

We would expect the infection intensity to show a similar geographical distribution pattern as the infection prevalence. Indeed, in both the 2016 and 2017 studies, we observed a positive association between infection prevalence and intensity at the village level. Similar observations have been made in other endemic settings for *S. mansoni* [58] and other *Schistosoma* species [67]. In general, variable transmission intensity is thought to account for these observations [68].

In 2017, we observed an overall infection prevalence of *S. mansoni* infection of 73.1%; almost double of 2016 (40.0%). Certainly, the different sampling procedure might account for the higher prevalence rate, as we purposely focused on known *S. mansoni* endemic villages. However, in 2017 we also employed a more sensitive diagnostic approach, consisting of an examination of two stools (4 Kato–Katz smears) and a urine sample (POC–CCA rapid test) per study participant. The infection prevalence ranged from 52.7% to 95.0% at health district level and from 32.3% to 96.2% at village level.

The Kato–Katz technique remains the standard method for diagnosing *S. mansoni*, however it has some shortcomings. It has a low sensitivity; it is time consuming and it requires skilled and trained technicians to identify the *S. mansoni* eggs microscopically. In addition, its sensitivity decreases with decreasing infection intensity [69]. The recently developed POC–CCA test [45] offers an alternative method. Its use in our study increased the number of *S. mansoni* patients identified. Of the 707 participants screened for *S. mansoni* using both techniques, 389 (55.0%) tested positive using the Kato–Katz technique and 442 (62.5%) tested positive using POC–CCA. Upon combining the results from the two techniques, 517 (73.1%) participants were diagnosed with *S. mansoni* infections. These observations are corroborated by the results reported by Okoyo and colleagues [70] when comparing the performance of CCA and KK techniques for evaluating *S. mansoni* infection in areas with low prevalence in Kenya. They found that using the CCA technique increased diagnostic accuracy. There is some discrepancy when comparing our results with those of Standley and colleagues [71], who found that CCA and KK techniques had a similar degree of accuracy. Recent publications discuss the specificity of POC–CCA. POC–CCA showed some cross-reactivity with intestinal nematodes and other health conditions, and therefore might overestimate the prevalence [72, 73].

The 2017 in–depth study was primarily conducted to assess the most important risk factors for an *S. mansoni* infection. We examined the demographic, socioeconomic, environmental, behavioural and knowledge risk factors. Among the most important risk factors, our multivariable logistic regression analysis identified socioeconomic factors, such as poor housing; environmental factors, such as living in a risky health districts, in close proximity to water bodies for long time periods; behavioural factors, such as the lack of a latrine, and swimming and washing in waterbodies; and knowledge factors [47].

In our study, gender was not a risk factor for infection as females and males had similar levels of infection prevalence in both studies, with a difference <2.0% in 2017 and >5.0% in 2016. These results are consistent with those of other authors [74, 75]. In Ituri province, men and women have different types of water contact activities, but contact intensities are comparable.

Age was an important risk factor. *S. mansoni* infection prevalence and intensity follow a typical age peak curve. In both studies, the prevalence and intensity increased with age until it peaked among the groups aged 10-19 years. Interestingly, the infection prevalence peak was higher in 2017 (86.1% *vs* 50.2% in 2016), but infection intensity was lower that year (45 epg *vs* 245 epg in 2016). Indeed, the use of a higher sensitivity diagnostic approach in 2017 is mainly responsible for these observations. Children of these age groups are most prone to have excessive mobility that may expose them to infected water while swimming, playing, bathing, washing clothes or fetching water [76]. Our results resemble the patterns reported by Kabatereine and colleagues [77] when describing the epidemiology of *S. mansoni* infections on the Ugandan side of Lake Albert, and by Tukahebwa and colleagues [74] when investigating *S. mansoni* infection in a fishing community on the shores of Lake Victoria.

A striking finding of our study is that children under five years are highly infected with *S. mansoni* (62.3% in 2017). This demonstrate the early life exposure of young children through bathing and playing in infested waters. A similar finding was reported by Nalugwa and colleagues [78], who described the high *S. mansoni* infection prevalence among preschool children in communities along Lake Victoria in Uganda.

Some health districts presented a higher risk of *S. mansoni* infection than others. *S. mansoni* is a focal parasite, meaning the more time people spend in affected areas, the more they are likely to get infected [52, 79].

Some positive associations were likely linked to both behavioural and environmental covariates, such as swimming and washing clothes in streams, which are widely practiced in the province and result in exposure to safe water.

Our study has some limitations. First, the sampling techniques during the two studies were not the same. In 2016, by sampling the first arrivals in the centre of the village, a selection bias may have occurred. Likewise, the population, especially in rural areas, is generally reluctant about research. They adhere more easily to interventions than to prior investigations. Thus, the study compliance in some villages was quite low. Men, in particular, adhered less strictly to the procedures. This difficulty resulted in fewer study participants in some villages. Second, we did not use the same diagnostic approach in each of the two study years. Also, the POC-CCA test was partially used in 2017 and we did not include molecular tests such as polymerase chain reaction (PCR) and/or loop-mediated isothermal amplification (LAMP) that were not available at the time in the region. Due to the prevailing insecurity due to armed groups in the province in 2016, we could only spend a maximum of two days in each village. Therefore, only one stool sample could be collected from each study participant. Given the low sensitivity of the diagnostic approach employed in 2016, we underestimated the true infection rates. Third, the risk factor analysis should be interpreted with caution, as the in–depth study did not include health districts and villages from the northern or hilly regions. Hence, some relevant risk factors might be missing. Fourth, we did not simultaneously conduct a malacological survey to assess the prevalence and rates of cercarial infection of snails in the region. This may have deprived us of valuable information on the transmission of the disease and its seasonality.

The DRC’s national NTD control program had completed mapping schistosomiasis and STH in 99.2% of the country’s health districts in 2015. It had launched a master plan 2016-2020 that work toward eliminating schistosomiasis as a public health problem by 2025, following the WHO guidelines. These guidelines depend on the level of infection as assessed in school age children (SAC). The subsequent recommendations are that preventive-chemotherapy (PCT) mass drug administration (MDA) be administered: i) annually for SAC and high–risk adults (HRA) in highly endemic communities (prevalence >50.0% in SAC); ii) every-other-year for SAC and selected high–risk adults in moderate to high prevalence communities (10-49.0%); iii) for SAC just twice during their primary school years in communities where prevalence is low (<10.0% in SAC) [5]. Another objective of the DRC NTDs program is to interrupt transmission by increasing access to adequate sanitation and drinking water and by improving the immediate environment of communities [35]. In Ituri province, control activities are performed by the provincial NTD branches situated in Aru and Bunia, which implement the control activities in the northern and southern health districts, respectively. To the best of our knowledge, only three health districts in the shore of Lake albert, namely Tchomia, Angumu, and Nyarambe, are benefiting the first strategy and all the others use the latter strategy of treating or have not started PCT yet. As the country is vast and without roads or trains, the supply of medicines is a major headache. Health districts routinely obtain their supplies from neighbouring Uganda. Even if the seasonality of transmission were not investigated, the best times for treatment could be during the two annual dry periods, December to February and June to July. During these periods, the weather is warm, and water becomes more and more scarce. People, especially children, make greater use of water courses for play, domestic, recreational, and occupational activities which expose them to infection. Our data will be of practical value to help improve control interventions, especially as COVID-19 became currently a huge global problem. COVID-19. Just to emphasize that this virus will surely make the work to be done even more complicated, not to mention all the people that will get ill and die.

The results of this study are important in several respects. They show that (i) *S. mansoni* is highly prevalent in Ituri province. The infection rates observed are beyond those reported in the 1960s [52] and those described in other parts of the country [13, 15, 20, 23, 24]. The observed infection levels are comparable with those in Ugandan fishing villages [74, 75]. The results also show that (ii) the age distribution of infection prevalence and intensity follows the typical pattern of an *S. mansoni* endemic area, where control measures are insufficiently implemented; and that (iii) preschool children bear a high infection burden and, therefore deserve special attention in the control programme.

In conclusion, our results provide comprehensive baseline data showing that *S. mansoni* is highly endemic and is a major health concern in Ituri province, DRC. Infection prevalence and intensity, and its relationship with the prevailing socioeconomic, environmental, and behavioural risk factors reflect intense exposure and alarming transmission rates. Our findings call for more robust plan of action for the control and, in the future, elimination of *S. mansoni* infection in the Ituri province. Intervention strategies could include the full implementation of WHO recommendations for mass drug administration (MDA) by treating communities adequately according to their real need. Furthermore, additional efforts are required to strengthen and expand community–based programs that promote practices aimed at preventing the spread of *S. mansoni* and other parasitic infections. Comprehensive community–based health education, and implementation of water, hygiene, and sanitation (WASH) programs both in rural and urban areas are of high value, especially in this context of the currently huge global problem of COVID-19 pandemic that will make the work to be done even more complicated, not to mention all the people that will get ill and die. Altogether, these efforts are likely to yield appreciable and sustainable gains in improving health and welfare of the population of Ituri province.

## Data Availability

Data will be prepared for availability when all the manuscripts based on the datasets have been submitted.

## Acknowledgements

We are grateful to all the study participants, both in 2016 and in 2017. Our sincere thanks to the research teams during this two–year period. We thank the Provincial Health Division officers, all the Health District officers, and the Institut Supérieur des Techniques Médicales de Nyankunde staff for their support, as well as the local authorities for their support during fieldwork.

## Supporting information captions

**Tables**

**Table S1**. *Schistosoma mansoni* infection prevalence and intensity in the 2016 geographical distribution study. Study conducted in 46 villages in Ituri province (n=2,131). One stool sample from each study participant was examined with the Kato–Katz test (two smears per stool).

**Table S2a**. *Schistosoma mansoni* infection prevalence and intensity in the 2017 in–depth study. Study conducted in 12 purposively–selected villages in Ituri province (n=707). Study participants provided two stool samples. From each sample, two Kato–Katz (KK) smears were examined (total four smears per person). In addition, each participant provided a urine sample for point–of–care circulating cathodic antigen (POC–CCA) test. Kato–Katz + POC–CCA combined results

**Table S2b**. *Schistosoma mansoni* infection prevalence and intensity in the 2017 in–depth study. Study conducted in 12 purposively–selected villages in Ituri province (n=707). Study participants provided two stool samples. From each sample, two Kato–Katz (KK) smears were examined (total four smears per person). Kato–Katz test results alone.

**Table S2c**. *Schistosoma mansoni* infection prevalence and intensity in the 2017 in–depth study. Study conducted in 12 purposively–selected villages in Ituri province (n=707). Study participants provided two stool samples. From each sample, two Kato–Katz (KK) smears were examined (total four smears per person). In addition, each participant provided a urine sample for point–of–care circulating cathodic antigen (POC–CCA) test. POC–CCA results alone (for prevalence).

**Table S3a**. Demographic and socioeconomic risk factors for *Schistosoma mansoni* infection (2017 in–depth study). Results obtained with the univariate analysis of risk groups for infection with *S. mansoni* among participants from 12 villages in Ituri province in 2017 (n=707).

**Table S3b**. Environmental risk factors for *Schistosoma mansoni* infection (2017 in–depth study). Results of the univariate analysis of risk groups for infection with *S. mansoni* among participants from 12 villages in Ituri province (n=707).

**Table S3c**. Behavioural risk factors for *Schistosoma mansoni* infection (2017 in–depth study). Results of the univariate analysis of risk groups for infection with *Schistosoma mansoni* among participants from 12 villages in Ituri province (n=707).

**Table S3d**. Family and individual risk factors for *Schistosoma mansoni* infection (2017 in–depth study). Results of the univariate analysis of risk groups for infection with *Schistosoma mansoni* among participants from 12 villages in Ituri province (n=707).

**Table S4**. Risk factors for *Schistosoma mansoni* infection (2017 in–depth study). Results of the multivariable analysis of risk groups for infection with *Schistosoma mansoni* among participants from 12 villages in Ituri province (n=707).

**Figures**

**Figure S1:** Geographic characteristics of Ituri province: This figure shows the study site: 1) Great geographic variability: high altitude up to 2,300 m above sea level and lowlands at about 500 m above sea level, large plateau and wide plains 2) Vegetation: dense forest in the southwestern region and grassy savanna along the eastern part 3) Water distribution and roads: a heavily irrigated region with several rivers, streams, swamps, ponds, and Lake Albert. There are very few roads made of laterite – the only passable road during the year is that which leads to Kibali gold mine in the neighbouring province of Haut-Uélé; only 5km of road are paved throughout the province, at Bunia city.

**Figure S2a:** Administrative subdivision of Ituri province: Ituri province is divided into 1) 5 administrative counties (or “territoires”): Mambasa (36,785 km^2^), Irumu (8,183 km^2^), Djugu (8,730 km^2^), Mahagi (5,216 km^2^), and Aru (6,749 km^2^); only 9% of Ituri’s population live in the largely forest-covered and reserve territory of Mambasa [39].

**Figure S2b:** Health districts in Ituri province: Ituri province is divided into 36 health districts: 14 were visited (brownish colour) during our two surveys in 2016 and 2017, including Adi, Laybo, Logo, Nyarambe, Angumu, Rethy, Bambu, Bunia, Tchomia, Nyankunde, Komanda, Lolwa, Mandima, and Nia-Nia [39].

**Figure S3:** Ethnic composition and population density in Ituri province: this figure shows that 1) Ituri province is populated by more than 40 tribal groups, belonging to one of five ethnic groups: Sudanese, Nilotic, Bantu, Nilo-Hamite, and Pygmy; 2) The mean density is about 80.8 inhabitants/km2. However, around 91.7% of the inhabitants live in 44.0% of the area, in the eastern half of the province, and only 8.3% live in 56.0% of the area, in the southwestern half of the province covered by the forest reserve [39].

